# Primary open angle glaucoma is associated with cortico-cortical receptive fields changes in early visual cortex

**DOI:** 10.1101/2025.01.03.25319969

**Authors:** Azzurra Invernizzi, Joana C. Carvalho, Joana Martins, Nomdo M. Jansonius, Remco J. Renken, Frans W. Cornelissen

## Abstract

**PURPOSE:** Primary open angle glaucoma (POAG, hereafter referred to as glaucoma) is a neuro-ophthalmic disease characterized by progressive degeneration of the retinal ganglion cells and nerve fibers. However, the exact pathogenesis of this disease remains unresolved, notably including its effect on the neural circuits of the visual pathway and cortex.

**METHODS:** We used both resting state (RS) and stimulus-driven functional MRI and applied a novel analysis technique (Bayesian Connective Field Modelling) to investigate the intra-cortical functional connective organization of the early visual cortex of patients with POAG. Analogous to population receptive field modelling for stimulus-driven activity, our connective field approach models how the activity in one cortical area (e.g. V2) can be explained based on that of another (e.g. V1).

**RESULTS:** We compared the CF parameters obtained for the early visual cortical areas in glaucoma to those of control participants. Our results show that in both RS and stimulus-driven conditions, CF sizes in early visual areas are smaller in glaucoma compared to control participants. To assess if these differences could be related to the ocular damage altering the visual input to the visual cortex, the control participants also observed the visual stimuli with a simulated scotoma (SS), designed to match the visual sensitivity of a participant affected by glaucoma as assessed using standard automated perimetry (SAP). In this condition, no differences in CF size were observed. Moreover, we found that CF size did not correlate with glaucoma severity, as assessed using both SAP and optical coherence tomography (OCT).

**CONCLUSION:** The observed differences in CF metrics may be the result of local reorganization or neurodegeneration of the early visual cortex that must have developed already at an early disease stage.

## 1. Introduction

Glaucoma is a neurodegenerative ophthalmic disease, which is characterized, amongst others, by loss of retinal ganglion cells and nerve fiber loss and a reduction in visual field (VF) sensitivity that typically starts in the periphery of the visual field. It is one of the most common causes of irreversible blindness worldwide. Recent brain imaging studies have shown that this degenerative damage is not limited to the eye and optic nerve but extends intracranially along the visual pathways and into the human visual cortex^1,2^. However, the extent to which this also affects the functioning of the adult visual cortex is not fully understood. The ability to establish this, as well as the presence of cortical adaptation (neuroplasticity) will be critical to assess the success of upcoming restorative therapies based on e.g. stem cells or cortical implants. Establishing the presence of neuroplasticity is challenging in eye diseases such as glaucoma since the visual input is already disrupted at the level of the eye. Such changes in visual input may result in spurious plasticity^3–5^. Potentially, this issue may be resolved by using resting-state (RS) fMRI recordings in combination with neural modelling of cortical functional connectivity. RS-fMRI assesses spontaneous fluctuations in cortical activity to estimate activation and connectivity patterns across the visual cortex areas at rest. It does so by leveraging the difference in the hemodynamic properties of oxygenated versus deoxygenated blood. These spontaneous fluctuations may indicate intrinsic alterations to the underlying neural circuitry of the brain, without relying on stimulation. Thus far, a few studies have used rs-fMRI to assess local functional connectivity in glaucoma data^6–11^, yet none of them have examined differences in local processing in the early visual cortical areas. An approach to specifically examine local spatial integration across brain areas is Connective Field (CF) modeling^12–14^. It translates the concept of the stimulus-driven receptive field (RF) into the neural connectivity domain. The^13^ approach assesses the spatial dependency between signals in distinct cortical visual areas. The CF is also known as the cortico-cortical population receptive field. Assessing CF properties based on RS-fMRI enables us to investigate cortical plasticity in glaucoma the absence of impoverished stimulus-related input, thus removing this potential confound in studies of the brain in eye diseases^12,14^.

We specifically used our novel Bayesian CF modeling^14,15^ approach to estimate CF properties (position, size and beta). Using both visual field mapping (VFM) and RS-fMRI, we investigate how glaucoma affects the functional organization of the visual cortex. To assess if these differences could be related to any short-term effects of the retinal loss affecting the visual input to the visual cortex, each of the control participants also observed the visual stimuli with a unique simulated scotoma (SS). This SS consisted of a mask on the display screen that was designed to mimic the reduction in visual sensitivity of a participant with glaucoma as assessed using standard automated perimetry (SAP).

Next, we compared the CF modelling results in these various experimental conditions to establish whether any observed changes in the CFs are related to glaucoma. To establish a potential relationship to disease severity, we correlated the CF properties to disease parameters as assessed using SAP and optical coherence tomography (OCT). Based on behavioral observations on spatial integration and visual crowding^16–18^, we expected to find increased CF sizes, with the increase being larger in cases with more severe glaucoma.

## 2. Methods

### 2.1 Study population

Nineteen patients with primary open-angle glaucoma (POAG) and 22 healthy participants were recruited. One POAG participant dropped out of the study, therefore only 18 participants were included in the analysis reported on in this manuscript. All participants were aged 60-75 years and satisfied all eligibility criteria for MRI scanning (i.e., no prior history of traumatic brain injury, having a pacemaker or being claustrophobic). Each participant underwent an ophthalmic screening (for details, see next paragraph) followed by two MRI sessions. Table 1 lists the population demographics and ophthalmic screening outcomes. The study procedures followed the Declaration of Helsinki and were approved by the ethics board of the University Medical Center Groningen (UMCG). Informed consent was signed by all participants prior to enrollment after all study procedures had been fully explained. One control participant was assigned to each participant with glaucoma, therefore only 18 controls were included in the final analysis. This pairing was done based on the participants’ age and gender.

**Table 1.** Demographics and ophthalmic characterization of the participants with glaucoma and controls.

| Measures | Participants with POAG (n = 18) |  | Controls (n = 18) |  |
| --- | --- | --- | --- | --- |
|  | Average | Standard Deviation | Average | Standard Deviation |
| Age | 69.6 | 8.7 | 68.2 | 8.41 |
| Gender (F%) | 48% | - | 34% | - |
| VA(R/L; decimal) | 1/0.9 | 0.4/0.3 | 1.1/1 | 0.2/0.4 |
| IOP(R/L; mmHg) | 13.5/13.6 | 2.6/3.7 | 12.9/13.2 | 3.1/3.4 |
| pRNFL thickness(R/L; $\mu\text{m}$) | 70.2/65 | 11.6/9.6 | 97/97.6 | 7.9/7.5 |
| VFMD (R/L; dB) | 6.7/ 8.7 | 8.1/8.3 | 0.3/0.1 | 1.5/1.3 |

Average and standard deviation of age, percentage of female, intraocular pressure (IOP) for the right and left eye (average over three measurements), average peripapillary retinal nerve fiber layer (pRNFL) thickness and the VF mean deviation (VFMD) measured with HFA for the right and left eye. Note that all POAG participants were receiving treatment when recruited to this study.

Inclusion criteria for the participants with glaucoma were as follows: intraocular pressure (IOP) > 21 mmHg before treatment onset; presence of a VF defect (glaucoma hemifield test outside normal limits) due to glaucoma in at least one eye; abnormal OCT (peripapillary retinal nerve fiber layer thickness [pRNFL] at least one clock hour P<0.01), and spherical equivalent refraction within ±3D. Exclusion criteria for both groups were: having any ophthalmic disorder affecting visual acuity (VA) or VF (other than POAG in the participants with glaucoma); any neurologic or psychiatric disorder; the presence of any abnormality or lesions in their magnetic resonance imaging (MRI) scans.

### 2.2 Ophthalmic assessment

Prior to their participation in the MRI experiments, both glaucoma patients and controls underwent an ophthalmic screening, which included the assessment of: visual acuity (VA); intraocular pressure (IOP); Humphrey Visual Field Analyzer (HFA) performed using a Carl Zeiss Meditec, Jena, Germany; and, structural assessment of the inner retina using a Canon OCT-HS100; Canon, Tokyo, Japan. Visual acuity was measured using a Snellen chart with optimal correction provided for the viewing distance. IOP was measured using a Tonoref non-contact tonometer (Nidek, Hiroishi, Japan). For HFA we used the 24-2 grid with the Swedish Interactive Threshold Algorithm (SITA) Fast. Only reliable HFA tests were included in this study; a visual field test result was considered unreliable if false-positive errors exceeded 10% or if both fixation losses exceeded 20% and false-negative errors exceeded^19^ 10%. Results of the structural assessment were expressed as the mean peripapillary retinal nerve fiber layer (pRNFL) thickness and the ganglion cell complex (GCC) thickness in the macular area.

### 2.3 Visual stimulus presentation and description

Visual stimuli were displayed on an MR compatible display screen (BOLD screen 24 LCD; Cambridge Research Systems, Cambridge, UK) located at the head-end of the MRI scanner. The participant viewed the screen through a mirror placed at 11 cm from the eyes, supported by the 32-channel SENSE head coil. The screen size was 22 x 14 degrees of visual angle and the distance from the participant’s eyes to the screen was 120 cm. The maximum stimulus radius was 7 deg of visual angle.

All participants underwent luminance contrast retinotopy, while the control participants also underwent luminance contrast retinotopy with simulated scotoma (SS) superimposed on the stimulus, as described in detail below. Stimuli were generated and displayed using the Psychtoolbox (https://github.com/Psychtoolbox-3/Psychtoolbox-3/) and VISTADISP toolbox (VISTA Lab, Stanford University), which are both MatLab based^20,21^.

#### 2.3.1 Luminance contrast retinotopy stimulus

All participants underwent binocular visual field (VF) mapping using a luminance contrast defined retinotopy (LCR) stimulus (Figure 1, panel A). The stimulus consisted of a drifting bar aperture (of 10.2 deg radius) with a high contrast checkerboard texture on a gray (mean luminance) background. A sequence of eight different bar apertures with four different bar orientations (horizontal, vertical, and two diagonal orientations), two opposite motion directions, and four periods of mean-luminance presentations completed the stimulus presentation, which lasted a total of 192 s per run. To maintain stable fixation, participants were instructed to focus on a small colored dot presented at the center of the screen and to press a button as soon as they noticed the dot changing color. Each participant performed four retinotopy scans.

**Figure 1.**
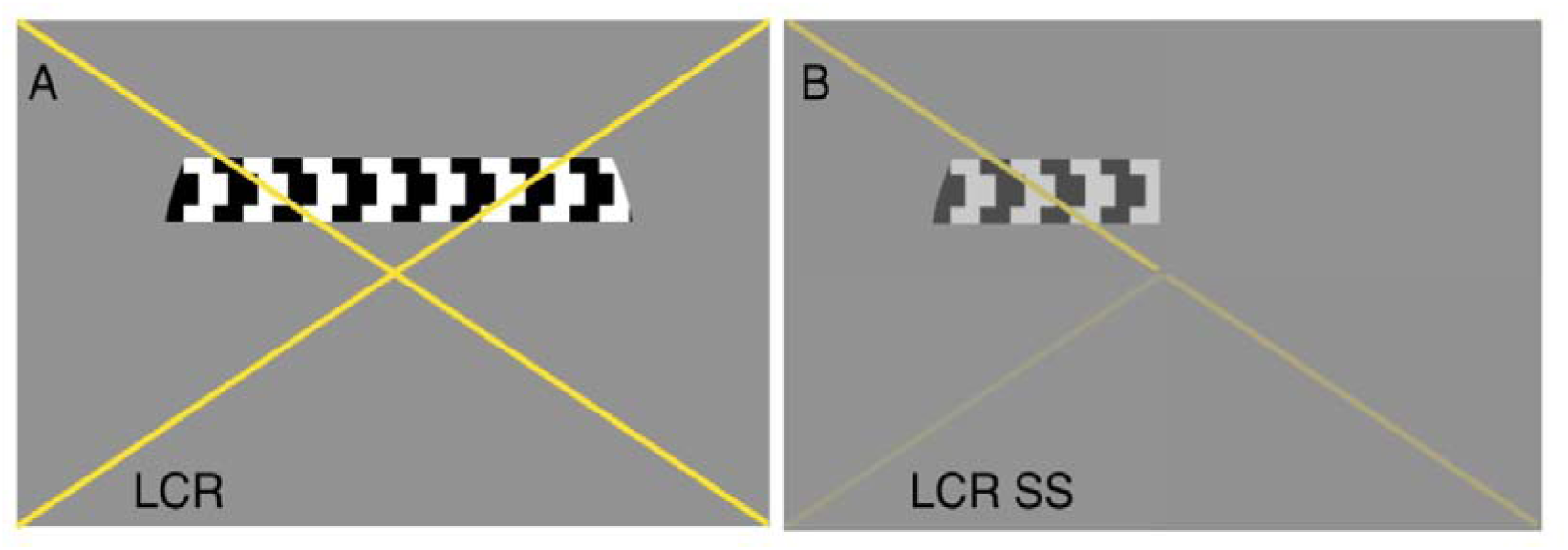
Example of the stimuli used to obtain pRF parameter estimates. Panel A shows LCR stimulus and panel B reports an example of LCR SS stimulus. This example depicts the contrast sensitivity loss of participant G10 (MD (OD)=-8.37 dB; MD(OS)=-14.72 dB). The color of the cross changed between yellow and black, and served to guide the gaze of the participant with central scotomas to the center of the cross.

#### 2.3.2 Luminance contrast retinotopy stimulus with a simulated scotoma superimposed

To control for the difference in visual perception between glaucoma and control participants, the control participants underwent an additional VF mapping scan during which they viewed the LCR stimulus with a simulated scotoma (SS) superimposed. The SS for a control participant was designed to simulate the contrast sensitivity of their matched glaucoma participant under binocular vision. The scotoma was simulated by masking the stimulus using local reductions in contrast applied through an alpha transparency contrast layer that was based on the HFA sensitivity values of the participant with glaucoma. For example, a decrease of 3dB in HFA sensitivity was simulated by means of a reduction in stimulus contrast of 50%. The simulated decrease in HFA sensitivity was determined by averaging, for each test location, the left and right eye dB value in the SAP total deviation plot of the glaucoma patient.

### 2.4 Resting-state fMRI

During the RS-fMRI scans, the screen was turned to black and the lights in the scanning room were turned off. All participants were instructed to keep their eyes closed, lay as still as possible, to not fall asleep and to try to not think of anything.

### 2.5 MRI and fMRI data acquisition procedure

Scanning sessions were performed on a 3 Tesla Siemens Prisma MRI-scanner using a 64-channel receiving head coil. In the first MRI session, an anatomical and two fMRI RS scans^3^ were acquired. For each participant, a T1-weighted scan (voxel size, 1mm; matrix size, 256 x 256 x 256) covering the whole-brain was recorded to chart cortical anatomy. Padding was used for a balance between comfort and reduction of head motion. The RS scans were collected using standard EPI sequence with duration of 260 seconds (TR, 1350 ms; TE, 30 ms;^3^ voxel size, 3mm, flip angle 68; matrix size, 94 x 94 x 45). Slices were oriented to be approximately parallel to the calcarine sulcus.

In the second MRI session, the retinotopic mapping took place. This session differed for participants with glaucoma and control participants. The control participants performed retinotopic mapping with both the regular LCR and the LCR with a simulated scotoma (LCR SS) (Figure 1). The functional scans were collected using a standard EPI sequence (TR, 1500^3^ ms; TE, 30 ms; voxel size, 3mm, flip angle 80; matrix size, 84 x 84 x 24). Slices were oriented to be approximately parallel to the calcarine sulcus. For all retinotopic scans (LCR and LCR SS, see section 2.2.4), a single run consisted of 136 functional images (duration of 204 s). The (S)SPZ localizers consisted of 144 functional images (duration of 216 s). Prior to the first retinotopy scan, a short T1-weighted anatomical scan with the same field of view as chosen for the functional scans was acquired. This scan facilitated the co-registration between the functional and anatomical volumes.

### 2.6 Data analyses

Pre-processing and analysis of fMRI data were done using ITKGray (http://www.itk.org),^22^ Freesurfer, and mrVista toolbox for MatLab environment (VISTASOFT, http://vistalab.stanford.edu/)^23–25^. The Bayesian CF approach^14,15^ was developed and implemented in MatLab 2016b (The Mathworks Inc., Natick, Massachusetts).

For each participant, structural, VFM, and RS functional MRI data were preprocessed. The structural scan was aligned in a common space defined using the anterior commissure-posterior commissure line (AC-PC line) as reference. Next, gray and white matter were automatically segmented using Freesurfer and manually adjusted using ITKGray (http://itk.org), in order to minimize segmentation errors. Then, all functional data were pre-processed using the mrVista toolbox. For both RS and VRM data the following steps are applied. First, head motion within and between scans were corrected by using robust multiresolution alignment of MRI brain volumes^26^, an alignment of functional data into anatomical space and an interpolation of functional data with segmented anatomical gray and white matter. For RS-fMRI data, a few additional denoising steps were applied. These steps were possible as the RS scans were acquired for the whole brain. First, spatial smoothing at 6mm FWHM was applied in order to perform the denoising step based on ICA-AROMA that identified noise and motion related components^27,28^. These components were then removed from the unsmoothed scans and then, RS-fMRI data were filtered by applying a band-pass filter with a high-pass discrete cosine transform with cut-off frequency of 0.01 Hz and a low-pass 4th order Butterworth filter with a cut-off frequency of 0.1 Hz.

#### 2.6.1 Population receptive field mapping

VFM scans were analyzed using a model-based analysis which allows localizing the visual field maps of interest, known as population receptive field (pRF) mapping^29^. In this analysis, for each voxel a pRF model (a 2D Gaussian) is convolved with the stimulus aperture and taking the hemodynamic response (HRF) into account, used to predict the BOLD response. Based on the best model fits, we estimate the visual field mapping parameters (eccentricity, polar angle and pRF size) per voxel. The parameters were projected onto a smoothed 3D mesh of the cortex. Using this analysis, the functional responses to the LCR stimulus were analyzed using a full field (FF) stimulus model. The data acquired in the LCR-SS condition in the control participants were analyzed using both this FF model and a model that included the SS. Based on the pRF data, the visual areas (V1, V2 and V3) were manually delineated to act as the source (V1) or the target region (V2, V3) in the subsequent CF analyses.

#### 2.6.2 Bayesian Connective Field mapping

CF models aim to explain the time-series for each location in a target region (V2 or V3) based^13^ on a weighted linear combination of the time-series in the source region (V1). The CF model is a 2D symmetric Gaussian kernel with the parameters CF position and CF size. For both VFM and RS data, we estimated the CF models using a Bayesian approach involving a Markov Chain Monte Carlo (MCMC) approach to efficiently sample the source region. The CF parameters associated with the best fitting model are converted from cortical units (cortical position) into visual field units (eccentricity and polar angle). This was done by inferring the^13^ pRF properties of the center voxel in the source region for each target location. During a total of 17500 iterations, CF models were computed of which the results of the first 10% of iterations were discarded to allow burn-in^30^. The posterior probability distributions of the CF parameters were estimated based on the remaining samples. The estimated model values were projected on a smoothed 3D mesh.

### 2.7 Spatial analysis

For evaluating the pRF properties, only voxels were included with an explained variance (VE) > 30% and an eccentricity > 1 deg and < 7 deg, resulting in 60% of voxels surviving. For evaluating the CF properties, only voxels with an VE > 30% were included. These pRF and CF thresholds were chosen based on common practice in the current literature^31–34^. For each target voxel in V2 and V3, the position for their CF in visual field space was taken to be the pRF of the center voxel of the weighted CF in the source region (V1).

To investigate differences in CF properties between glaucoma and control participants, a cumulative distribution of each CF parameter was estimated at the participant level. Furthermore, the effect size (beta) was retained as an additional CF parameter^15^. Group level statistical testing was performed by first calculating the median across voxels per participant, and secondly, comparing these median values across groups (glaucoma vs control participants, glaucoma vs control participants with SS and finally, control participants vs control participants with SS). Based on a quantile analysis of the posterior distribution^14,35^, we computed a voxel-wise uncertainty measure for each CF parameter by subtracting the upper (Q3) and lower (Q1) quantile of the posterior distribution. The estimated uncertainty was computed for both the RS and VFM data for each participant and repeated for each parameter directly estimated by the CF model (CF size and beta). Permutation analysis (10000 repetitions) with FWE correction applied for the number of group level comparisons was used. P-values <= 0.05 were considered significant. The correlation between CF size and disease severity was calculated using a linear fixed effects model, with a slope and intercept per participant as random effects. Finally, the intraclass correlation coefficient (ICC,^36,37^ was computed to estimate the test-retest reliability between the two RS-fMRI scans. Based on previous literature^14^, the 5% most active voxels based on VE were used to compute the ICC score.

## 3. Results

### 3.1 Cortical Topography

At the group level, no differences were found for any of the pRF parameters across the visual areas between the control participants (with and without SS) and the glaucoma participants (Figure S2). The complete analysis of the pRF parameters are reported in the supplementary materials.

Figure 2 shows, for an example participant couple, that for both the RS- and VFM-derived values, the topographical maps of the eccentricity and polar angle of the CF were comparable and showed a marked visuotopic organization in both the healthy and glaucoma participants. At the group level, only the CF size differed between glaucoma and healthy participants in both the VFM and RS conditions (Figure 2). CF maps for the second RS (RS2) scan are reported in supplementary material (Figure S1). Table 2 lists the median and interquartile ranges at the group level for each CF parameter. Notably, both glaucoma and age-matched healthy participants are characterized by small CF size compared to young healthy participants^35^.

**Figure 2.**
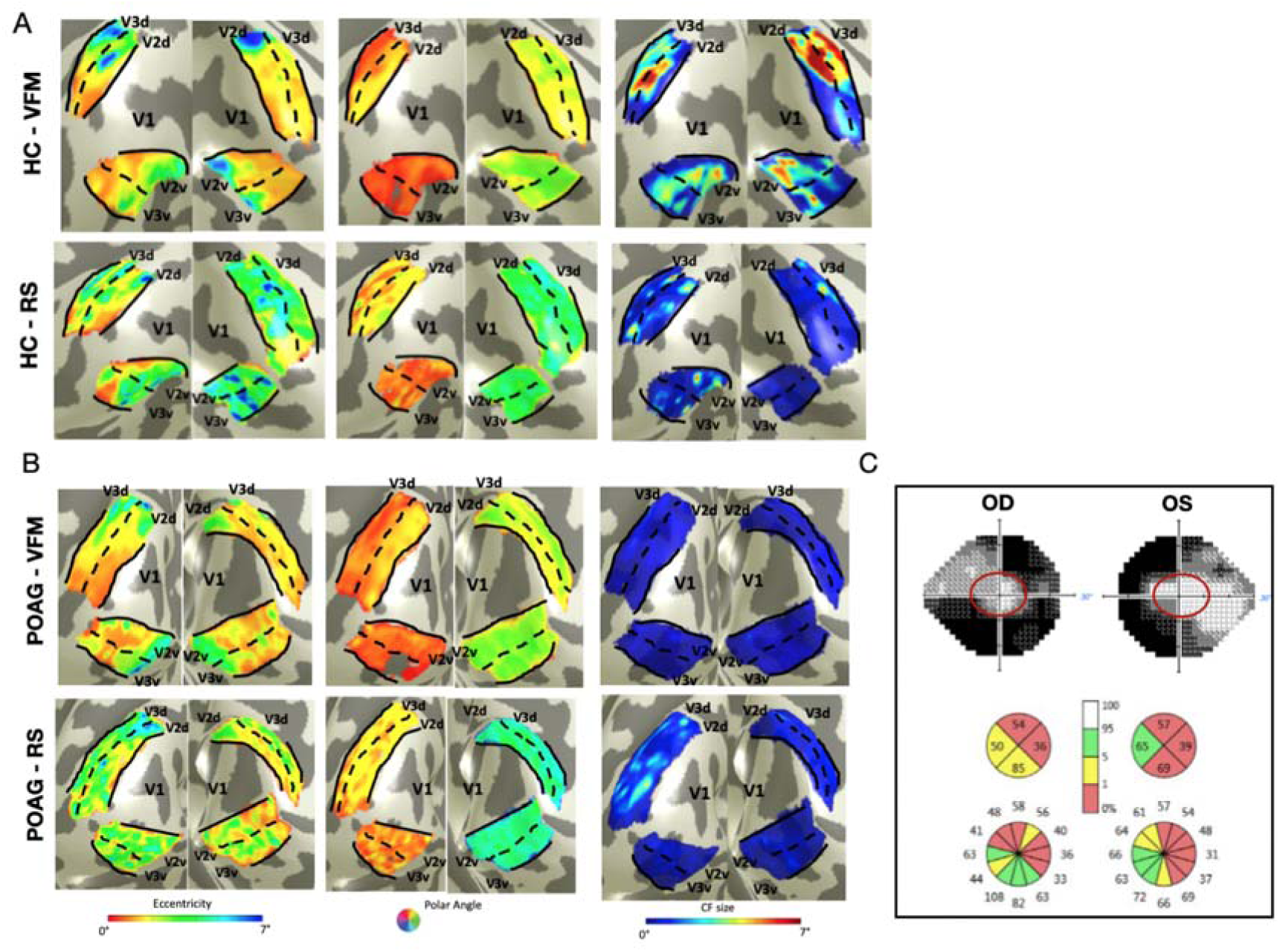
Connective Field topography based on VFM and RS-fMRI. Panels A and B show maps for a glaucoma participant and an age matched control participant, respectively. From left to right: eccentricity, polar angle and CF size maps obtained from VFM (upper rows) and RS (bottom rows). It is notable that in the glaucoma participant a good retinotopic organization is maintained. Panel C shows a graphic representation of the VF and OCT data for the glaucoma participant in panel B. Note that in the VFM experiment, the portion of the visual field inside the red circle (up to 7 deg eccentricity) was stimulated.

**Table 2.** Group median and interquartile range estimated based on VFM and RS data.

| V1>V2 | Control |  | Control SS |  | Glaucoma |  |
| --- | --- | --- | --- | --- | --- | --- |
|  | median | IQR [Q1, Q3] | median | IQR [Q1, Q3] | median | IQR [Q1, Q3] |
| Eccentricity | 3.633 | [2.11, 6.85] | 3.67 | [1.81, 5.95] | 2.26 | [1.46, 4.82] |
| Polar Angle | 2.55 | [1.12, 3.96] | 2.65 | [1.08, 4.02] | 2.48 | [1.77, 3.73] |
| CF size | 0.48 | [0.15, 1.43] | 0.52 | [0.15, 1.08] | 0.38 | [0.15, 0.56] |
| Beta | 1.18 | [0.91, 1.49] | 1.13 | [0.89, 1.37] | 1.35 | [1.01, 1.68] |
| Unc. CF size | 0.53 | [0.21, 1.04] | 0.25 | [0.16, 0.53] | 0.19 | [0.15, 0.27] |
| Unc. Beta | 0.09 | [0.04, 0.21] | 0.01 | [0.04, 0.19] | 0.01 | [0.04, 0.19] |
| V1>V3 | Control |  | Control SS |  | Glaucoma |  |
|  | median | IQR [Q1, Q3] | median | IQR [Q1, Q3] | median | IQR [Q1, Q3] |
| Eccentricity | 4.19 | [2.15, 6.62] | 4.01 | [1.963, 6.53] | 2.49 | [1.48, 4.34] |
| Polar Angle | 2.56 | [1.05, 4.03] | 2.82 | [1.08, 4.03] | 2.9 | [1.49, 3.78] |
| CF size | 0.82 | [0.21, 2.94] | 0.67 | [0.31, 1.48] | 0.47 | [0.17, 0.67] |
| Beta | 1.21 | [0.96, 1.47] | 1.18 | [0.93, 1.39] | 1.45 | [1.12, 1.78] |
| Unc. CF size | 0.91 | [0.31, 2.22] | 0.47 | [0.26, 1.51] | 0.22 | [0.15, 0.34] |
| Unc. Beta | 0.15 | [0.04, 0.21] | 0.01 | [0.04, 0.21] | 0.11 | [0.05, 0.22] |

RS1
| V1>V2 | Control |  | Glaucoma |  |
| --- | --- | --- | --- | --- |
|  | median | IQR [Q1, Q3] | median | IQR [Q1, Q3] |
| Eccentricity | 3.82 | [2.48, 6.89] | 3.15 | [1.90, 5.17] |
| Polar Angle | 2.67 | [1.42, 4.12] | 2.29 | [1.75, 3.57] |
| CF size | 0.71 | [0.21, 2.64] | 0.5 | [0.18, 0.76] |
| Beta | 1.45 | [0.99, 1.81] | 1.46 | [0.99, 1.87] |
| Unc. CF size | 0.29 | [0.18, 0.91] | 0.16 | [0.11, 0.24] |
| Unc. Beta | 0.11 | [0.05, 0.27] | 0.08 | [0.04, 0.16] |
| V1>V3 | Control |  | Glaucoma |  |
|  | median | IQR [Q1, Q3] | median | IQR [Q1, Q3] |
| Eccentricity | 4.02 | [2.60, 6.56] | 3.57 | [1.93, 5.28] |
| Polar Angle | 2.54 | [1.37, 3.95] | 2.58 | [1.38, 3.68] |
| CF size | 0.62 | [0.18, 2.04] | 0.51 | [0.18, 0.77] |
| Beta | 1.29 | [0.95, 1.71] | 1.41 | [1.0, 1.81] |
| Unc. CF size | 0.28 | [0.15, 1.13] | 0.17 | [0.11, 0.24] |
| Unc. Beta | 0.12 | [0.05, 0.28] | 0.09 | [0.04, 0.18] |

RS2
| V1>V2 | Control |  | Glaucoma |  |
| --- | --- | --- | --- | --- |
|  | median | IQR [Q1, Q3] | median | IQR [Q1, Q3] |
| Eccentricity | 3.81 | [2.48, 6.89] | 2.62 | [1.63, 4.84] |
| Polar Angle | 2.67 | [1.42, 4.11] | 2.25 | [1.64, 3.58] |
| CF size | 0.71 | [0.21, 2.64] | 0.48 | [0.17, 0.69] |
| Beta | 1.45 | [0.99, 1.81] | 1.7 | [1.13, 2.20] |
| Unc. CF size | 0.29 | [0.18, 0.91] | 0.16 | [0.11, 0.23] |
| Unc. Beta | 0.11 | [0.05, 0.27] | 0.07 | [0.03, 0.17] |
| V1>V3 | Control |  | Glaucoma |  |
|  | median | IQR [Q1, Q3] | median | IQR [Q1, Q3] |
| Eccentricity | 4.02 | [2.60, 6.56] | 3.16 | [1.88, 5.55] |
| Polar Angle | 2.54 | [1.37, 3.95] | 2.37 | [1.55, 3.47] |
| CF size | 0.62 | [0.18, 2.04] | 0.43 | [0.17, 0.65] |
| Beta | 1.29 | [0.95, 1.71] | 1.69 | [1.23, 2.12] |
| Unc. CF size | 0.28 | [0.15, 1.13] | 0.17 | [0.11, 0.24] |
| Unc. Beta | 0.12 | [0.05, 0.28] | 0.08 | [0.04, 0.21] |

For eccentricity, polar angle, CF size, beta and the estimated uncertainty, the median and interquartile were computed at the single participant level and then concatenated at the group level for each ROI separately.

### 3.2 Connective Field size

Figure 3 shows the cumulative distributions of CF size for glaucoma and control participants as obtained during VFM and both RS-fMRI conditions. For VFM, the CF size was significantly different between glaucoma and control participants (with and without SS) for both V1>V2 and V1>V3 (Table 3, VFM panel). For RS, the CF size differed significantly for V1>V2 in both RS scans, while for V1>V3 it reached significance in RS2 and bordered on it in RS1 (Table 3, panels for RS1 and RS2). Note that the individual cumulative distributions for each participant in RS2 scan are shown in Figure S3. Figure 4 shows the cumulative distributions of the uncertainty in the estimated CF sizes for glaucoma and control participants. The uncertainty in CF size differed between glaucoma and control participants in both the VFM and RS scans for both V1>V2 and V1>V3 (Table 3). Overall, this indicates that CF sizes in V1 are smaller for V2 and most likely also for V3 in participants with glaucoma compared to controls, with the uncertainty in the size estimates being overall smaller for the glaucoma compared to the control groups.

**Figure 3.**
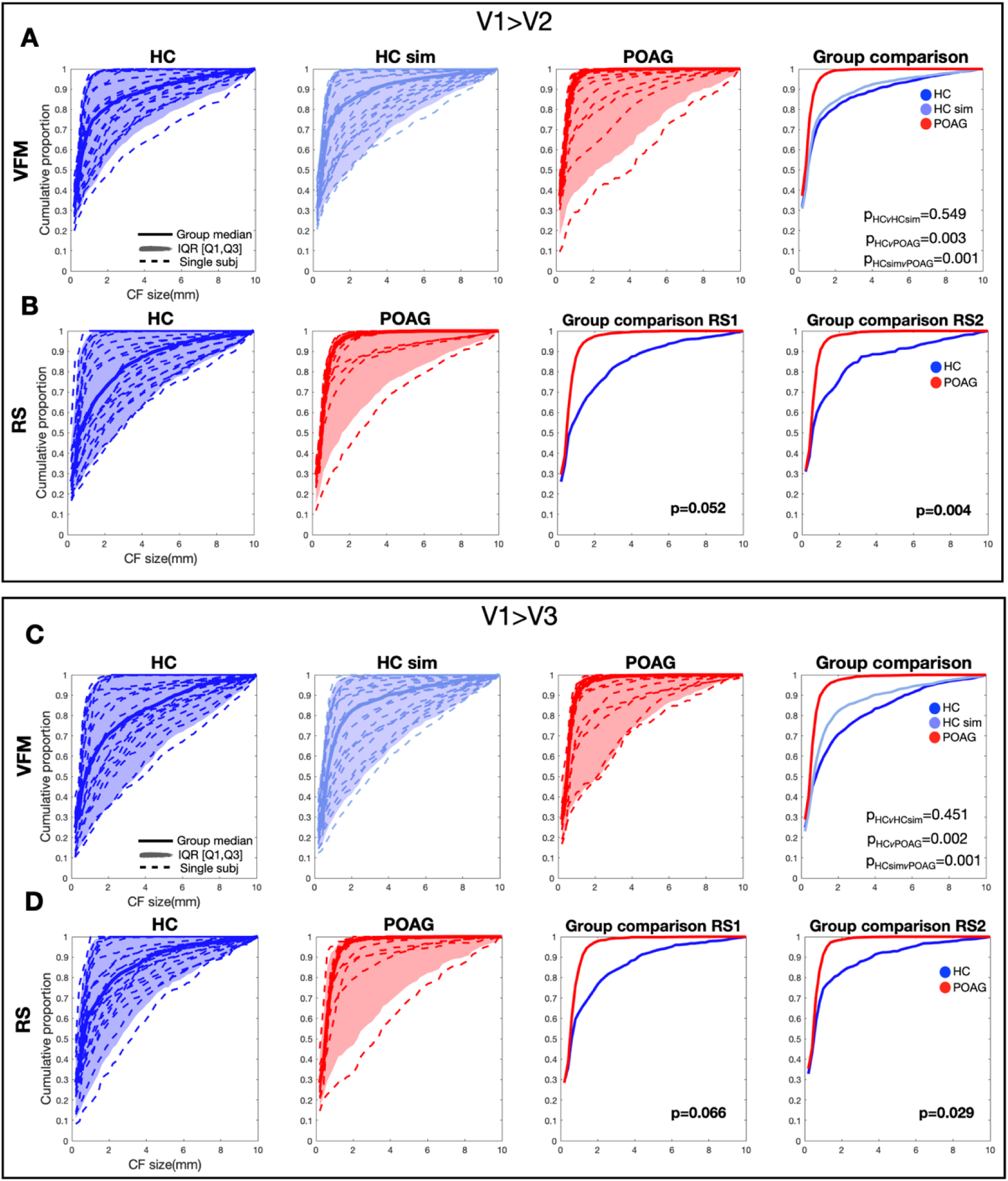
Cumulative distribution of connective field size at the group level based on VFM and RS fMRI data. The cumulative distribution of each individual participant (dashed line), the group median (dash line) and the interquartile range are plotted for participants with glaucoma (red), controls (dark blue) and controls with a simulated scotoma (light blue). Panels A and C represent the distributions based on VFM in the three groups, while Panels B and D do so for the RS scan 1. The final figure in Panels A and C compare group median distributions. The final two figures in Panels B and D compare the group median distributions obtained during RS scans (RS1 and RS2). The individual distributions obtained in RS scan 2 are reported in the supplementary material.

**Figure 4.**
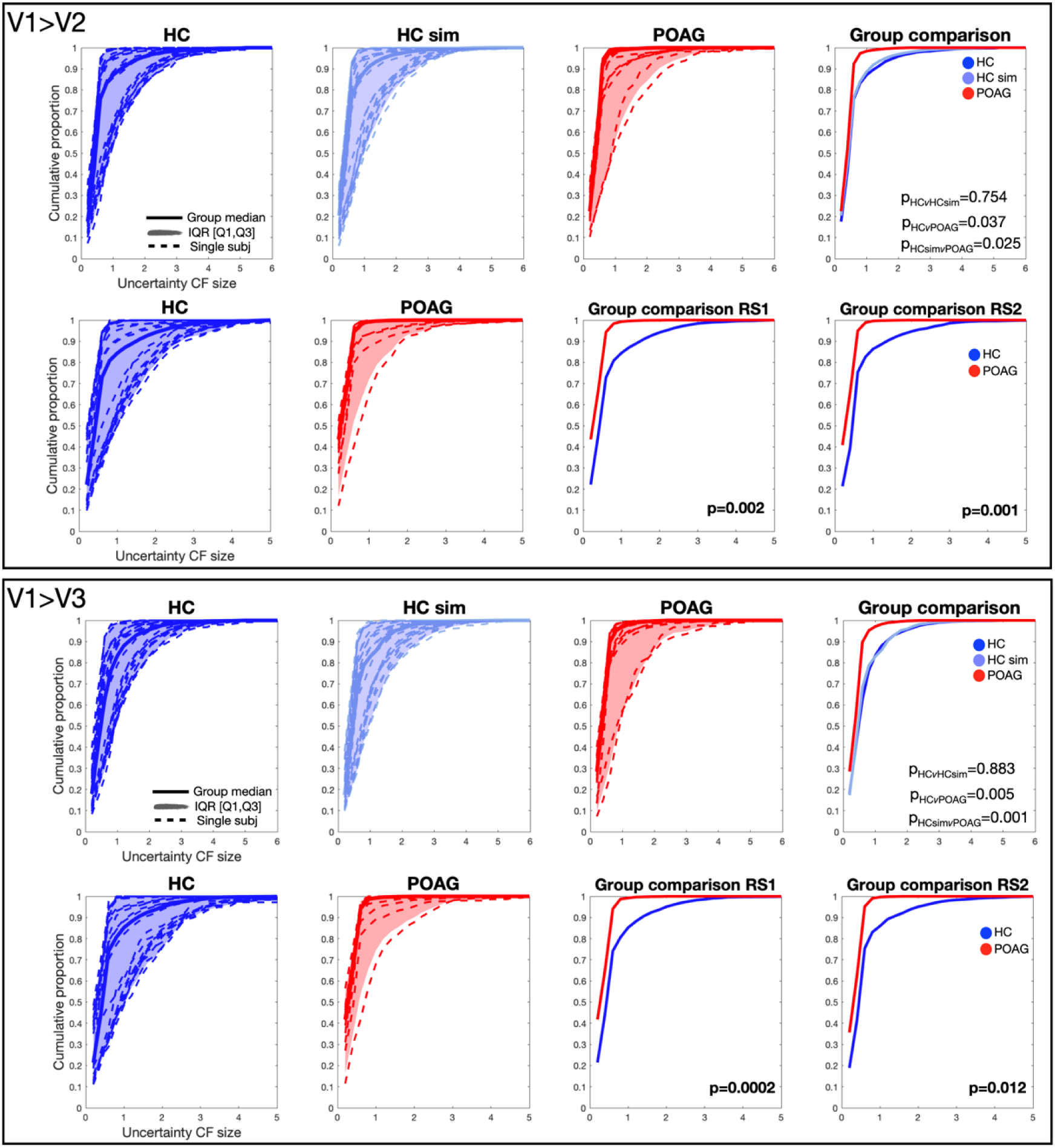
Cumulative distribution of uncertainty in the connective field size at group level based on VFM and resting-state data. The cumulative distribution of each individual participant (dashed line), the group median (dash line) and the interquartile range are plotted for participants with glaucoma (red), controls (dark blue) and controls with a simulated scotoma (light blue). Panels A and C represent the distributions based on VFM in the three groups, while Panels B and D do so for the RS scan 1. The final figure in Panels A and C compare group median distributions. The final two figures in Panels B and D compare the group median distributions obtained during RS scans (RS1 and RS2). The individual distributions obtained in RS scan 2 are reported in the supplementary material.

**Table 3.**
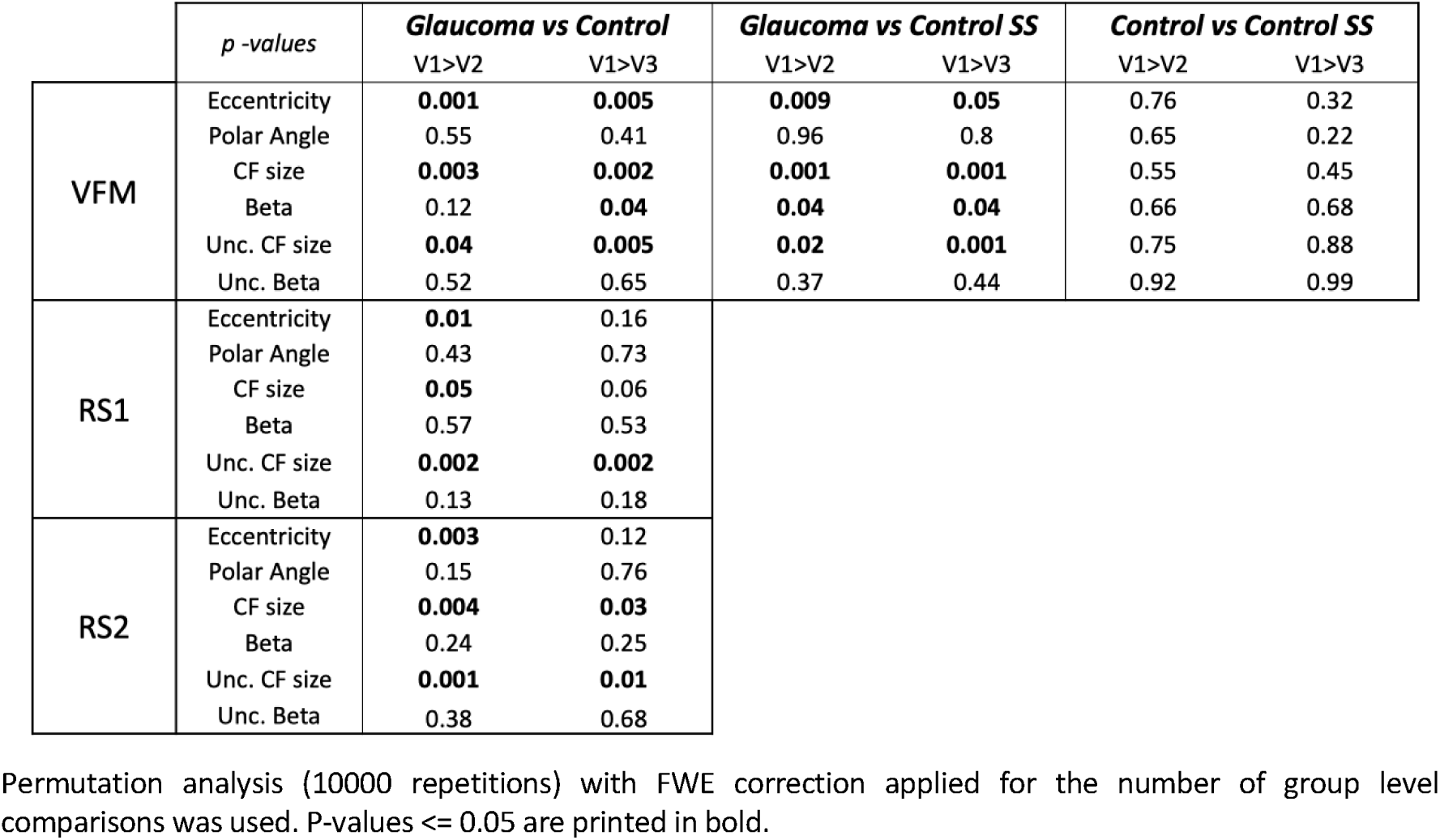
P-values for differences in group median in the VFM and RS conditions.

### 3.3 Connective Field beta

Figure 5 shows the cumulative distributions of beta (effect size) for glaucoma and control participants as obtained during both VFM and RS-fMRI. Only for the VFM data, for both V1>V2 and V1>V3 the differences between the glaucoma and control participants (with and without SS) reached significance. No significant differences were found for either of the two RS scans (Table 3, panels RS1 and RS2). Finally, Figure 6 shows the cumulative distribution in uncertainty for the estimates of beta in both glaucoma and control groups. No significant differences were found between groups for either the VFM or RS data (Table 3). Together, this indicates that the effect sizes were larger during VFM in glaucoma participants compared to both the standard and SS conditions, but not in the RS conditions. For both groups, the estimates were obtained with comparable uncertainty in all conditions.

**Figure 5.**
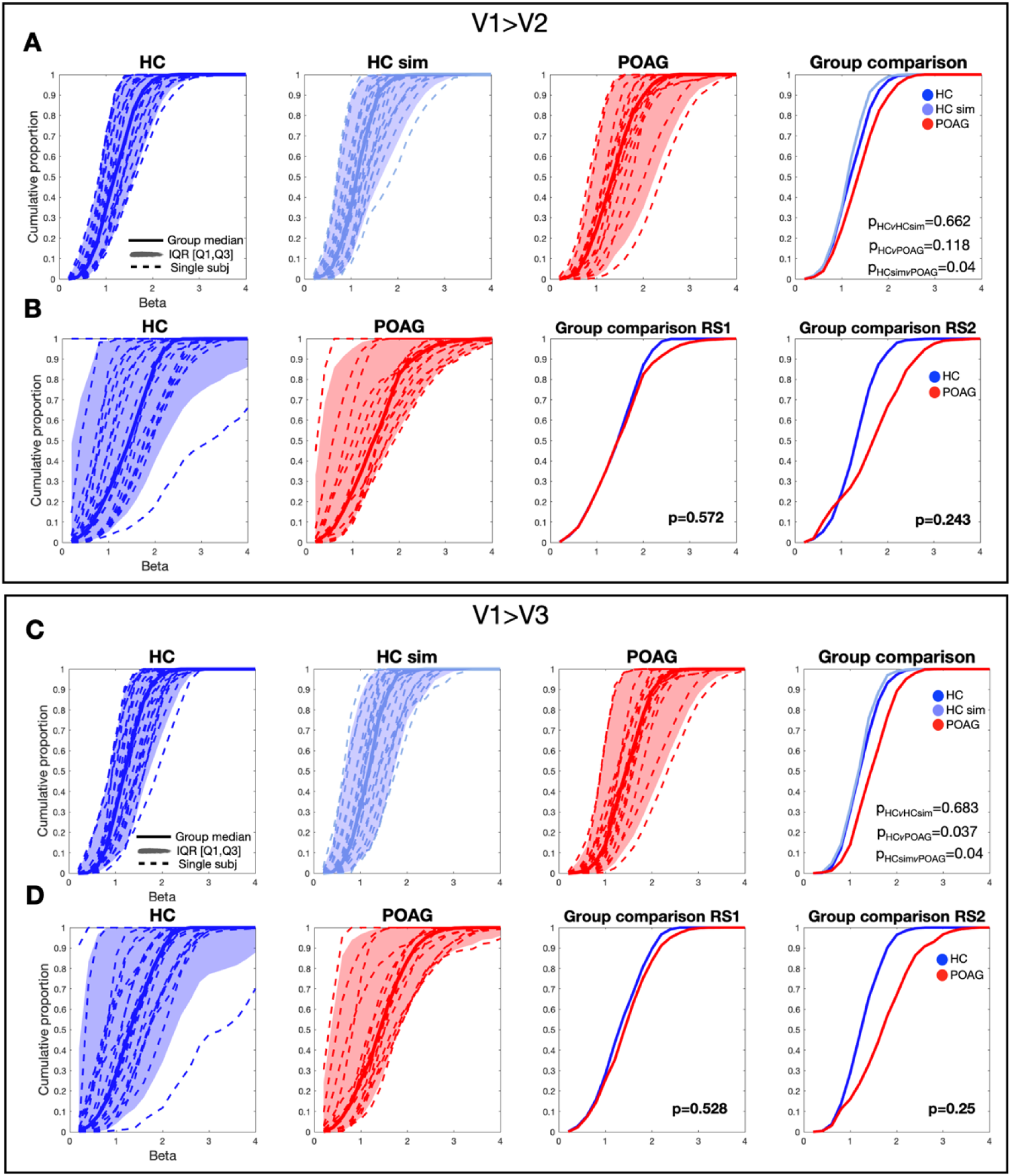
Cumulative distribution of beta at the group level based on VFM and resting-state data. The cumulative distribution of each individual participant (dashed line), the group median (dash line) and the interquartile range are plotted for participants with glaucoma (red), controls (dark blue) and controls with a simulated scotoma (light blue). Panels A and C represent the distributions based on VFM in the three groups, while Panels B and D do so for the RS scan 1. The final figure in Panels A and C compare group median distributions. The final two figures in Panels B and D compare the group median distributions obtained during RS scans (RS1 and RS2).

**Figure 6.**
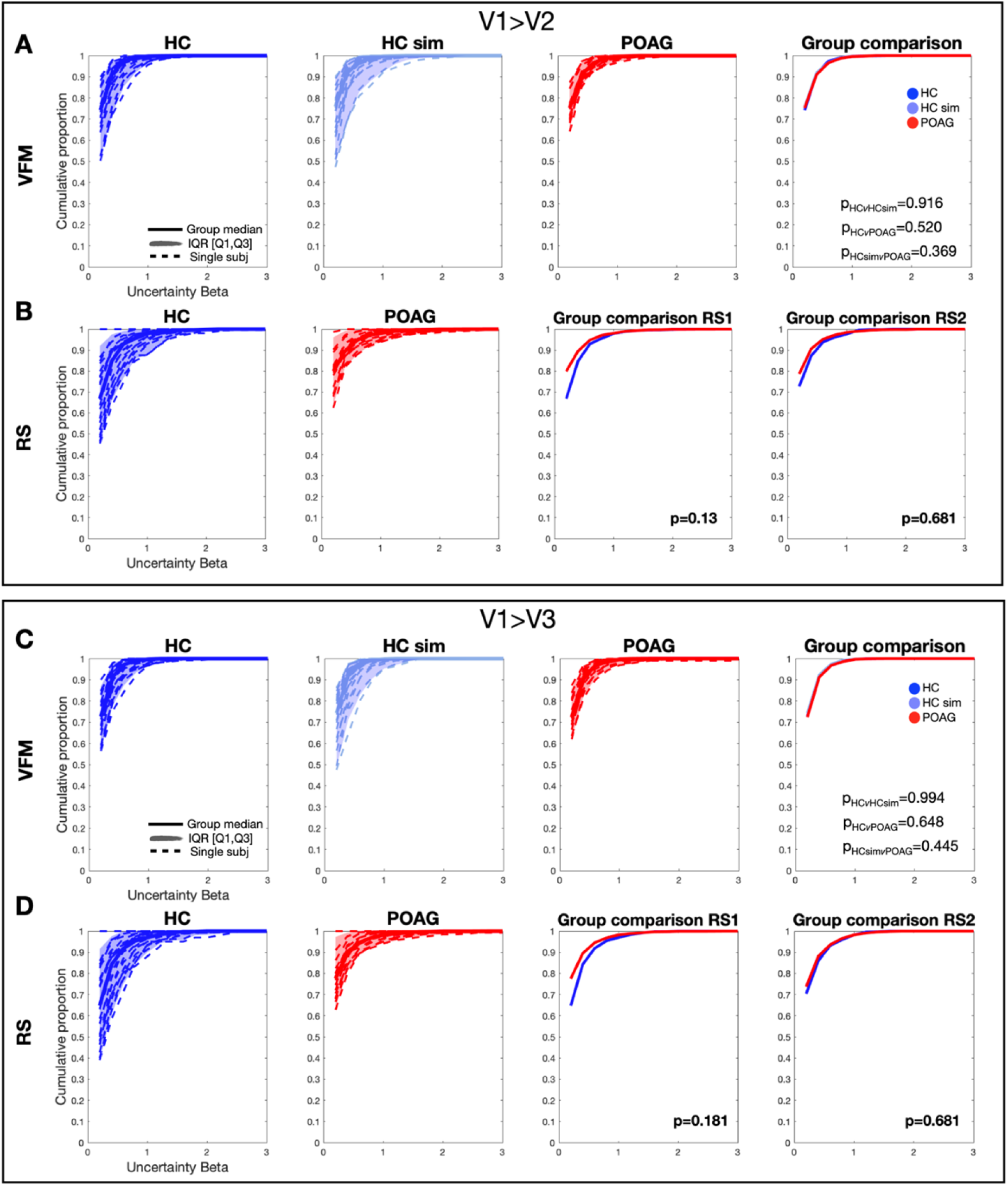
Cumulative distribution of the uncertainty in beta estimates obtained for the VFM and resting-state conditions. The cumulative distribution of each individual participant (dashed line), the group median (dash line) and the interquartile range are plotted for participants with glaucoma (red), controls (dark blue) and controls with a simulated scotoma (light blue). Panels A and C represent the distributions based on VFM in the three groups, while Panels B and D do so for the RS scan 1. The final figure in Panels A and C compare group median distributions. The final two figures in Panels B and D compare the group median distributions obtained during RS scans (RS1 and RS2).

### 3.4 Connective Field eccentricity

Figure 7 shows the cumulative distributions of CF eccentricity for glaucoma and control participants as obtained during both VFM and RS-fMRI. The difference between groups in both the VFM and RS conditions was significant for V1>V2 but not for V1>V3. The distributions of polar angle did not differ between groups for any visual area or experimental condition, for more details see Supplementary Material (VFM- or RS-based, Figure S4). This indicates that the CF of V2 in V1 had slightly lower eccentricity in the glaucoma participants compared to controls (in both the standard and SS conditions), while their polar angle was similar.

**Figure 7.**
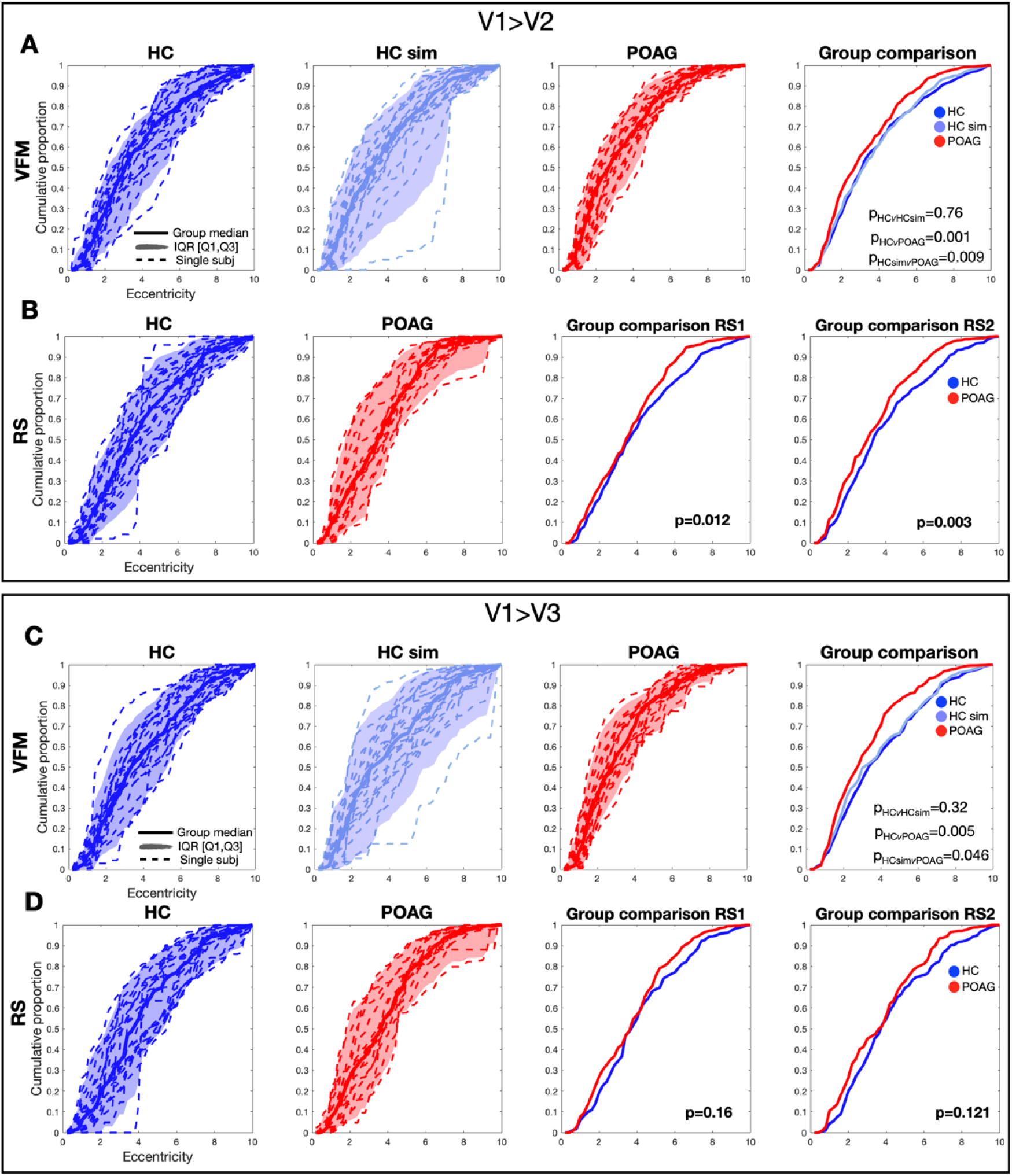
Cumulative distribution of CF eccentricity at the group level based on VFM- and RS fMRI data. The cumulative distribution of each individual participant (dashed line), the group median (dash line) and the interquartile range are plotted for participants with glaucoma (red), controls (dark blue) and controls with a simulated scotoma (light blue). Panels A and C represent the distributions based on VFM in the three groups, while Panels B and D do so for the RS scan 1. The final figure in Panels A and C compare group median distributions. The final two figures in Panels B and D compare the group median distributions obtained during RS scans (RS1 and RS2). The individual distributions obtained in RS scan 2 are reported in the supplementary material.

### 3.5 CF size as a function of glaucoma severity

Figure 8 shows the correlations between the average CF size in the individual visual field quadrants (< 7 degrees) and the sensitivity (MD) (panel A, top figures) or the OCT-based GCC thickness (panel A, bottom figures) in the corresponding quadrant. Panel B shows the correlations with the RS-derived CFs. For both control and glaucoma participants the CF size for V1>V2 exhibits weak and non-significant correlations. Only the moderate correlation between GCC-thickness and CF size in glaucoma participants borders on significance. For the V1>V3 CFs, the correlation plots are shown in the supplementary materials (Figure S6). Furthermore, we report the correlations based on RS2 data (Figure S7).

**Figure 8.**
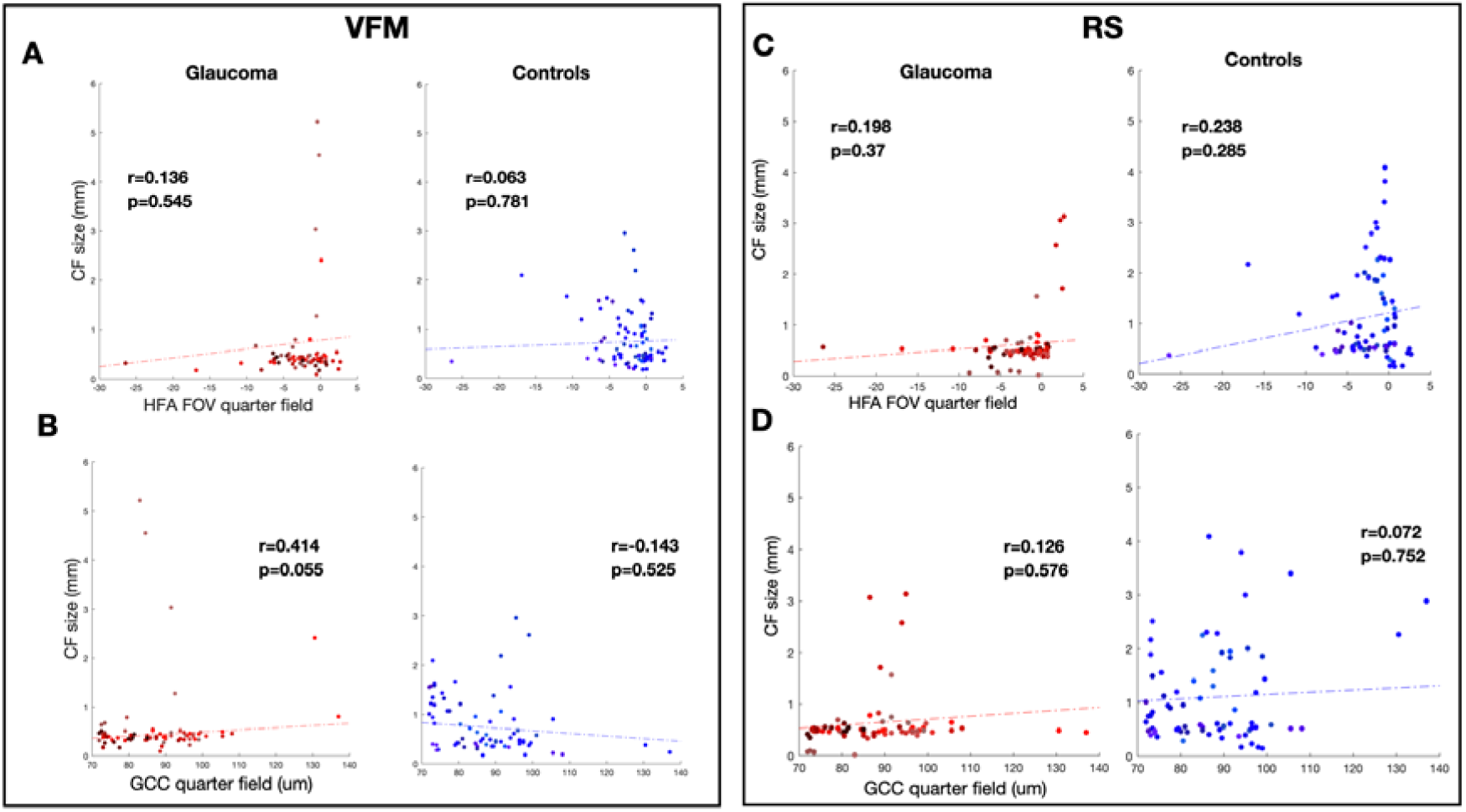
CF size for V1>V2 as a function of either HFA or OCT data of glaucomatous participants. Correlation of CF size VFM-based obtained from individual quadrants with HFA and GCC from healthy and glaucoma participants. HFA values were calculated as mean deviation of both eyes combined. GCC thickness was calculated by averaging the thickness of the macula of both eyes. Each data point is from a separate quadrant of an individual participant. Panels B and D show the correlations of VFM- and RS-based CF size with GCC value, respectively.

## 4. Discussion

Our main finding is that connective (intracortical receptive) fields in the early visual areas are, on average, smaller in glaucoma compared to control participants. This was the case in both VFM and RS conditions, indicating that this difference was found both in the presence and absence of visual stimulation. The CF approach models how the brain spatially integrates information between visual brain areas. Thus, our results suggest that the way in which the primary visual cortex integrates and propagates visual information to higher order visual areas may have changed as a result of glaucoma.

CF size did not have a marked relation to the severity of glaucoma, as assessed using SAP and optical coherence tomography (OCT). Although small shifts in CF eccentricity were observed in glaucoma participants, the coarse retinotopic organization of the early visual cortex in glaucoma patients is preserved, which is in line with previous findings in other pathology^31,38^. We conclude that differences in CF may be the result of local reorganization or neurodegeneration in the early visual cortex that may have started already at an early disease stage. Below, we discuss our results in more detail.

### 4.1 Glaucoma is characterized by reductions in CF size

Based on brain activity recorded during both visual stimulation and resting-state conditions, we found shifts in the position and reductions in the size of the CFs. The smaller CFs are not a mere consequence of ocular damage resulting in impoverished visual input reaching the brain in glaucoma. This can be concluded from our finding that simulating comparable scotoma as in the patients in the control group did not result in reduced CF sizes.

In addition, we applied Bayesian CF modeling^15^. This allowed us to also quantify the variability and reliability associated with each of the estimated CF parameters. No differences were observed in the estimates of effect size (beta) or in its associated uncertainty. This indicates that the observed differences are not due to low signal quality or a poor fit of the CF models to the data in either of the groups. Moreover, the reduced uncertainty in the CF size in glaucoma participants further supports this conclusion. Finally, our results were consistent over two resting-state scans (supplementary material, test-retest). Note that for both CF size and its associated uncertainty, a floor effect may have^15^ affected the results, due to a minimum CF size (i.e.0.01) enforced by the MCMC model. However, such influences are unlikely to be able to explain the substantial differences in the distributions in CF size that we observed.

### 4.2 Reductions in CF size are not related to glaucoma disease stage

We found no marked correlations between CF size and glaucoma severity (either in terms of VF sensitivity or thickness of the ganglion cell complex; Figure 6). Only a weak, borderline significant, relation between CF size and thickness of the ganglion cell complex was found in the VFM condition. This implies that the differences in CFs are not clearly related to the amount or quality of the signals that are passed on from the eyes to the brain. Moreover, glaucomatous visual field defects start out in the visual periphery, with the central visual field assumed to remain relatively unaffected by the disease. Due to the relatively limited field of view (FOV) of stimulation in the scanner (14 deg diameter), we could mostly evaluate the cortical projection zone of the fovea and parafovea. This further corroborates our notion that the observed differences in cortical visual information processing are not a mere consequence of glaucoma impoverishing the visual input to the brain. Although glaucomatous visual field defects show similarity in their progression, individual glaucoma patients’ visual field defects are unique. This is reflected in specific and unique patterns of reorganization at the brain level as observed with retinotopic mapping^39^. Our present study adds to this previous observation by showing that also the intracortical processing is affected, on average. The fairly large differences in the distributions of individual participants suggests that this aspect too may be unique related to their individual disease characteristics.

### 4.3 Compensatory mechanisms in glaucoma

Glaucoma is primarily characterized by a progressive loss of RGCs^40,41^. This loss of RGCs is reflected in mechanisms that may compensate for the structural-functional discordance present in glaucoma. An enlargement of Ricco’s area, which expresses the degree of spatial summation across the visual field, has been observed in glaucoma. Such an enlargement may serve as a compensatory mechanism for improving signal detection and contrast sensitivity^42–44^. Such an enlargement of Ricco’s area could be the result of increased signal pooling^45^ from the most responsive neurons and other neural compensatory mechanisms. In parallel, also increased levels of peripheral visual crowding in glaucoma point in the same direction^46^. Crowding is the result of spatial pooling of visuospatial information. The CF model has been conceived as a mathematical representation of such cortical (spatial) integration and pooling as the signal propagates from V1 to higher visual areas^13,35^.

Consequently, the enlargement of Ricco’s area and increased crowding would predict larger CFs in glaucoma, whereas we find smaller ones. At present, we see no obvious way in which to reconcile this contrast between our fMRI findings and previous behavioral results. A possible difference may come from the larger eccentricities at which the results on Ricco’s area and crowding have typically been obtained compared to the more central field assessed in our current fMRI study. One speculative explanation could be that the cortex tries to compensate for loss of resolution or attempts to ‘undo’ any increased integration at earlier processing levels. A further possibility is that the CF measures, through their reliance on the BOLD response, also reflect differences in cortical perfusion in glaucoma for which there is some evidence at the level of the cortex and retina^47–49^. Future studies could investigate this by integrally examining Ricco’s area, visual crowding, connectivity, and perfusion in early^40^ visual cortex.

### 4.3 Glaucoma as a neuro-ophthalmic disease

A progressive loss of RGC, optic disc damage and retinal nerve fiber layer changes have been widely reported in glaucoma^50^ and point towards the neurodegenerative nature of POAG. In the absence of a cure, current guidelines emphasize therapies for slowing down progression by lowering intraocular pressure (IOP), the most important and modifiable risk factor in POAG^51^. Over the last two decades, evidence has accumulated that indicates that the neural damage also extends intracranially and affects the entire visual pathway^52,53^. Glaucoma is also characterized by local cortical reorganization^54–60^.

Our findings and those of others suggest that connectivity changes in the brain are present already in the early stages of the disease. Indeed, the majority of the patients in our study had scotomas that were characterized by relatively small reductions in contrast sensitivity. Yet, the relatively large variability in CF properties amongst patients render it less obvious what could be gained from including fMRI in diagnostics. However, fMRI-based approaches may become relevant for characterizing the degree of neurodegeneration and neuroplasticity in individual patients with advanced glaucoma in the light of future therapeutic approaches, e.g. those based on stem-cells. The quality of the remaining visual pathways and cortical connectivity could be a biomarker for staging a therapy and in follow-up. In particular, an approach based on resting-state fMRI could be compatible with a clinical setting as it is non-invasive and demands little effort from patients.

### 4.4 Limitations and future directions

The limited field of view (FOV) of the visual stimulation in our study made us focus our present work on the central visual field of patients. Consequently, we could not evaluate connectivity in the visual cortex that represents the farther visual peripheral. By studying participants with more advanced glaucoma and scotomas with binocular overlap within the MRI FOV it could be investigated whether the observed differences in CF properties would be more profound in such cases.

Another possible limitation was the heterogeneity in the location and extent of the scotomas. This made it harder to average results over participants. To, at least partially, overcome this limitation, we correlated CF properties to MD and retinal thickness at the quarter field level. Moreover, it is likely that differences in disease duration and progression have also influenced results. To more deeply understand the origin of the CF differences, future studies may either study many more participants with glaucoma at various disease stages or small cohorts that are more homogenous in their characteristics.

We found similar CF properties in participants with glaucoma during VFM and RS fMRI. Therefore, to reduce participant burden, future studies could consider focussing exclusively on RS-fMRI and delineate visual brain areas using anatomy-based and probabilistic maps^61^. The information from such maps could potentially be used to also guide the Bayesian CF model in estimating local CF properties, e.g. based on an estimate of cortical magnification. Finally, for historical reasons, the MR acquisition protocols between our VFM and RS scans were slightly different in our present study. Nevertheless, the CF estimates obtained based on the RS and VFM data agreed well. Nevertheless, future studies should consider using identical protocols in the VFM and RS scans.

## 5. Conclusion

Glaucoma is characterized by smaller CF sizes in the early visual cortical areas. The fMRI-derived differences can be found during both RS and visual stimulation indicating these are not caused by impoverished visual stimulation as a result of the eye disease. Moreover, these do not show a marked relation with disease stage. This suggests these differences may be a result of neurodegeneration or cortical plasticity that already occurred at an early stage of the disease.

## Supplementary Material

### Test-Retest

To estimate test-retest reliability between the two RS scans, we selected the 5% most active voxels and computed the ICC scores for each CF parameter obtained from RS1 and RS2 scans. The arbitrary threshold level for CF estimates was chosen based on^14^. A positive ICC value is reported for all CF parameters and across all rois (Figure S5, V1>V2 - panel A; V1>V3 - panel B).

## Supplementary Figures

**Figure S1.**
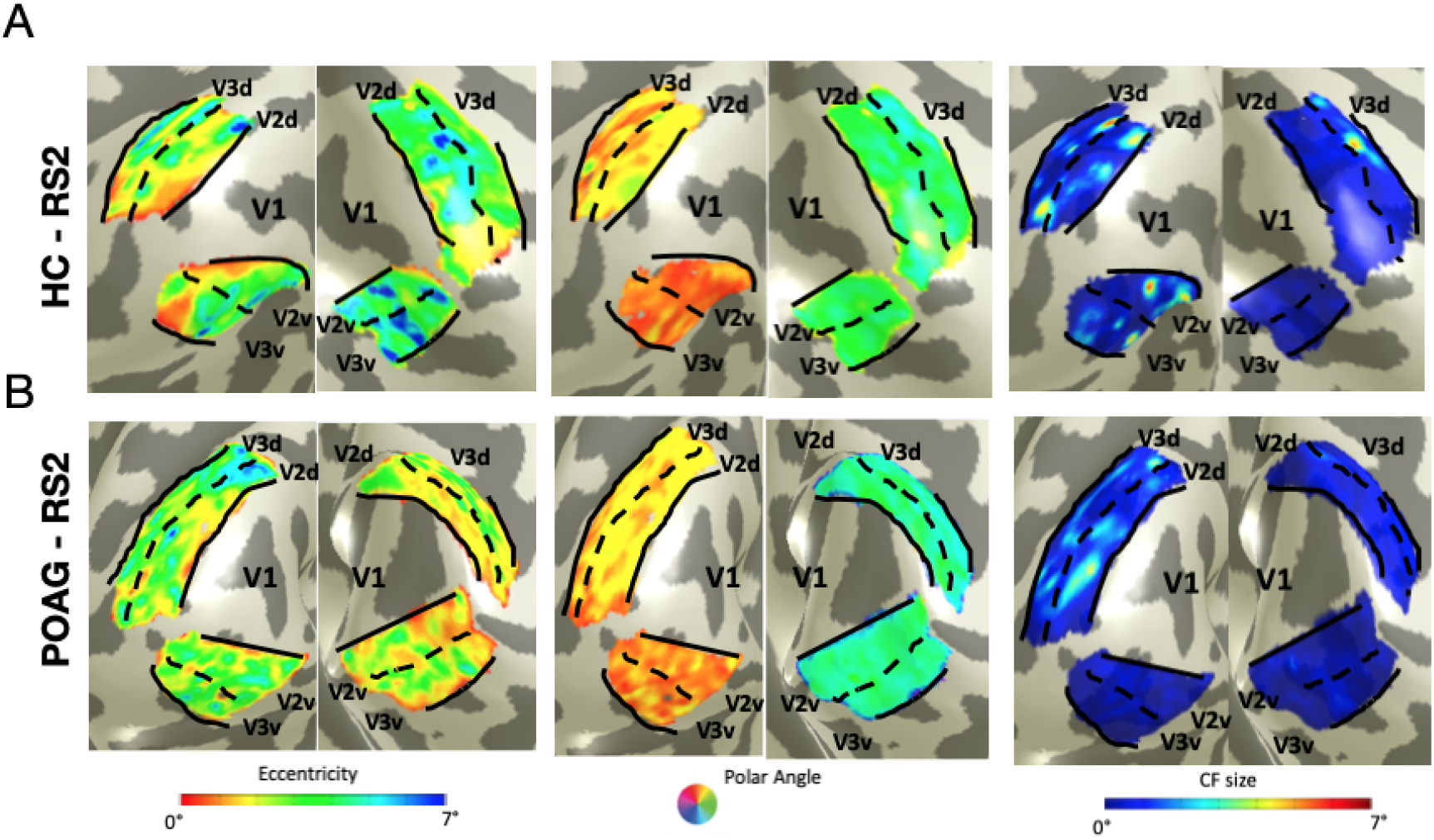
Visualization of CF maps based on RS2 scan for one control and one glaucoma participant. From left to right: eccentricity, polar angle and CF size parameters obtained from RS2 data. Panel A corresponds to an age-matched healthy participant, while Panel B to a glaucoma participant.

**Figure S2.**
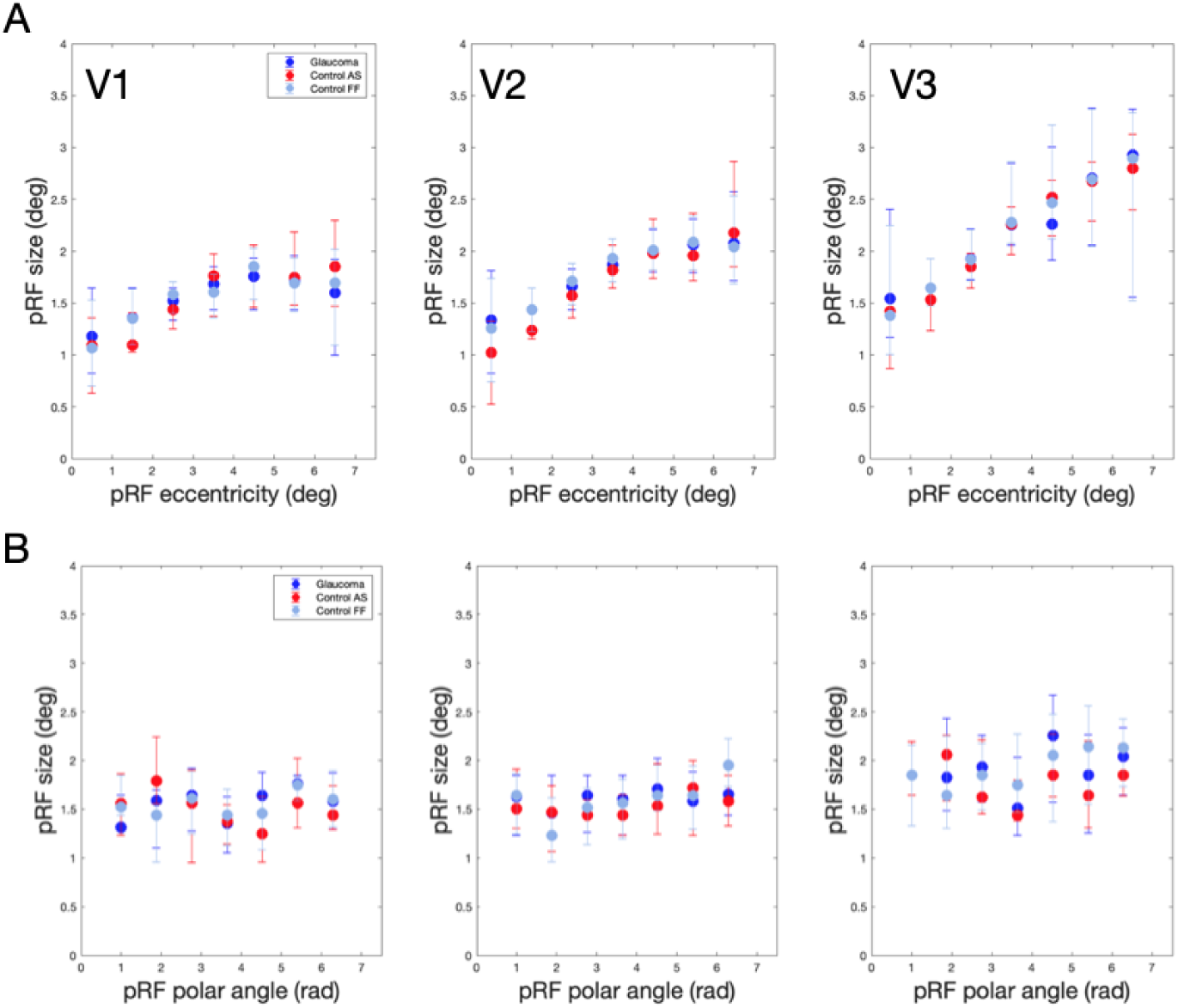
Average pRF properties in early visual areas V1, V2 and V2 as a function of pRF eccentricity (A) and polar angle (B). Eccentricity was binned in bins of 1 deg. Results for participants with glaucoma (red dot), control participants with (light blue dot) and without a simulated scotoma (blue dot). Error bars indicate the standard error of the mean over participants per hemisphere.

**Figure S3.**
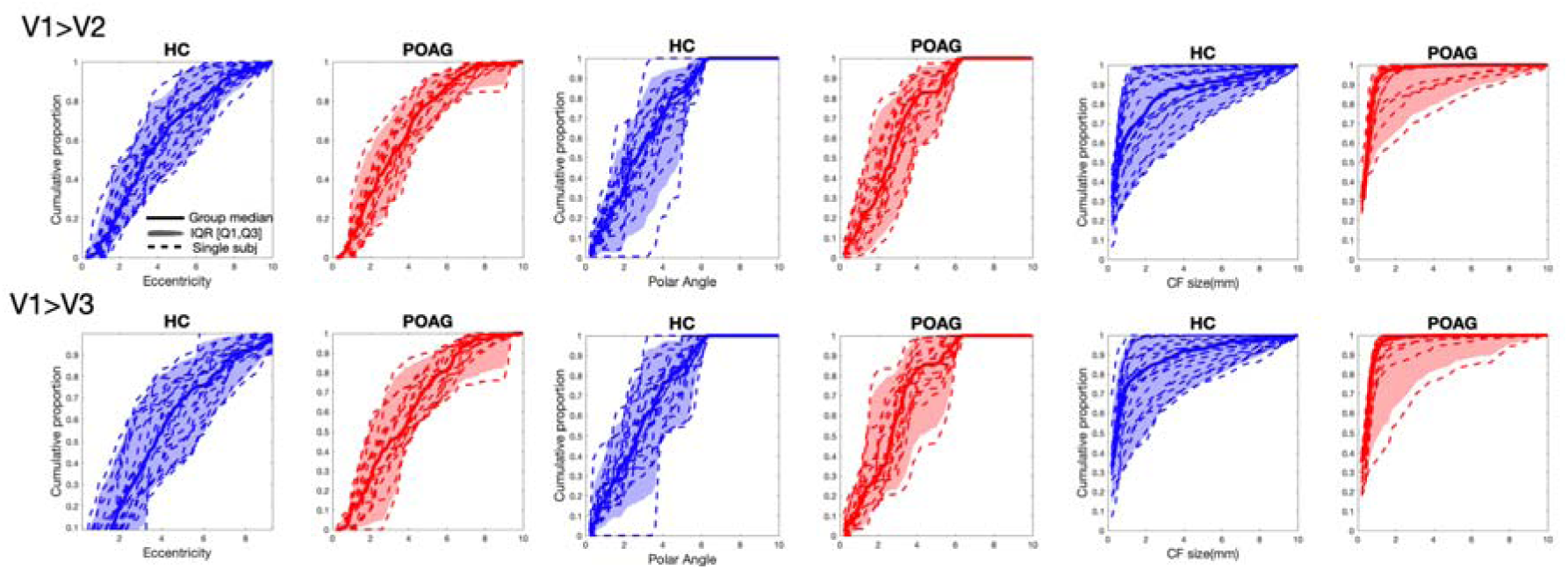
Cumulative distributions of CF parameters eccentricity, polar angle and size, based on data from the resting-state scan 2. The cumulative distribution of each individual participant (dashed lines), the group median (thick continuous line) and the interquartile range (shaded area) was plotted for participants with glaucoma (red) and control (blue).

**Figure S4.**
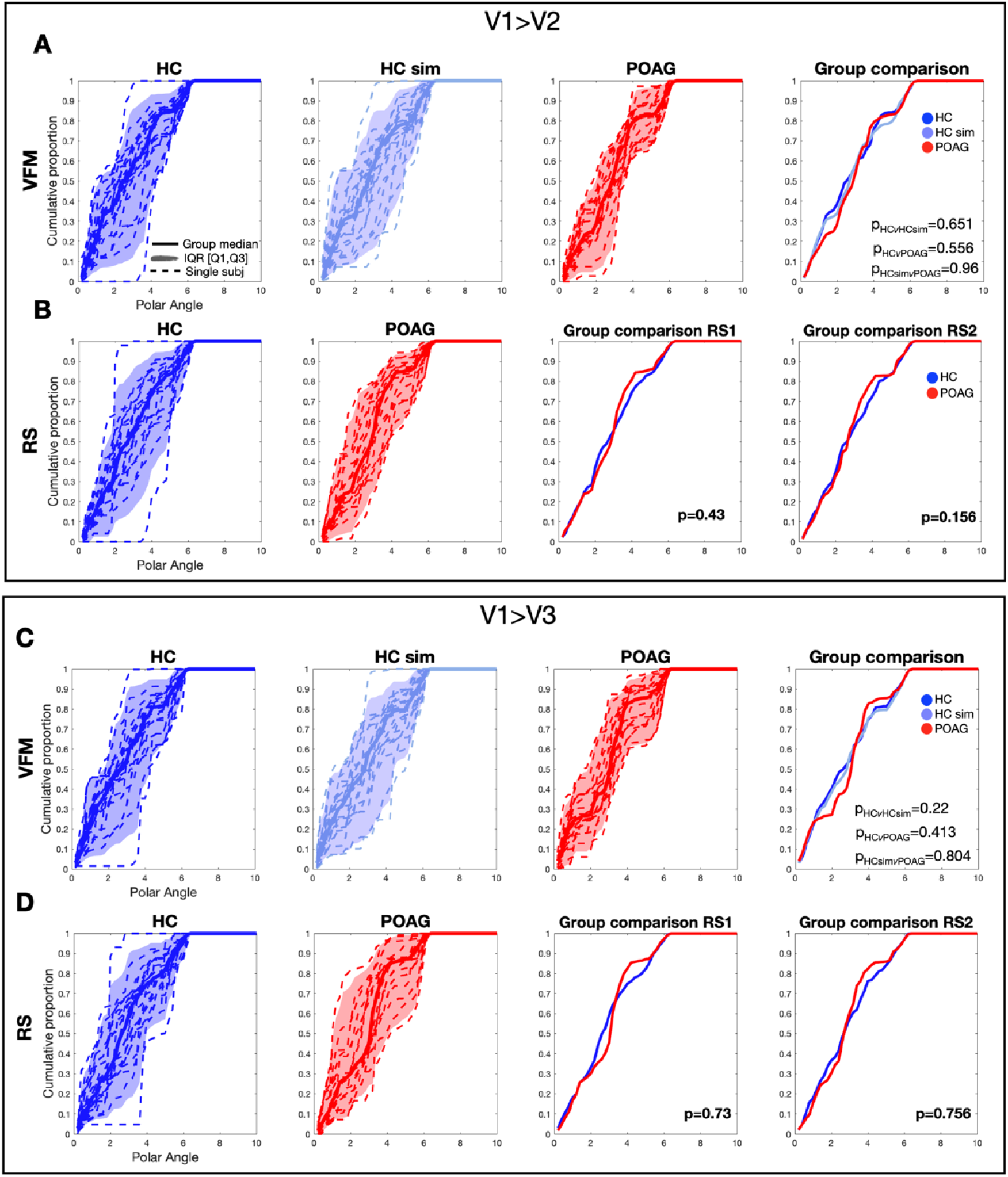
Cumulative distribution of polar angle based on VFM and resting-state data. The cumulative distribution of each individual participant (dotted line), group median (dash line) and the interquartile range are plotted for glaucoma (red), control (blue) and control with simulated scotoma (light blue). Panel A shows distributions for the VFM data in the three groups. Panel B shows the distributions obtained for RS1 and the median group results for both RS scans (RS1 and RS2).

**Figure S5.**
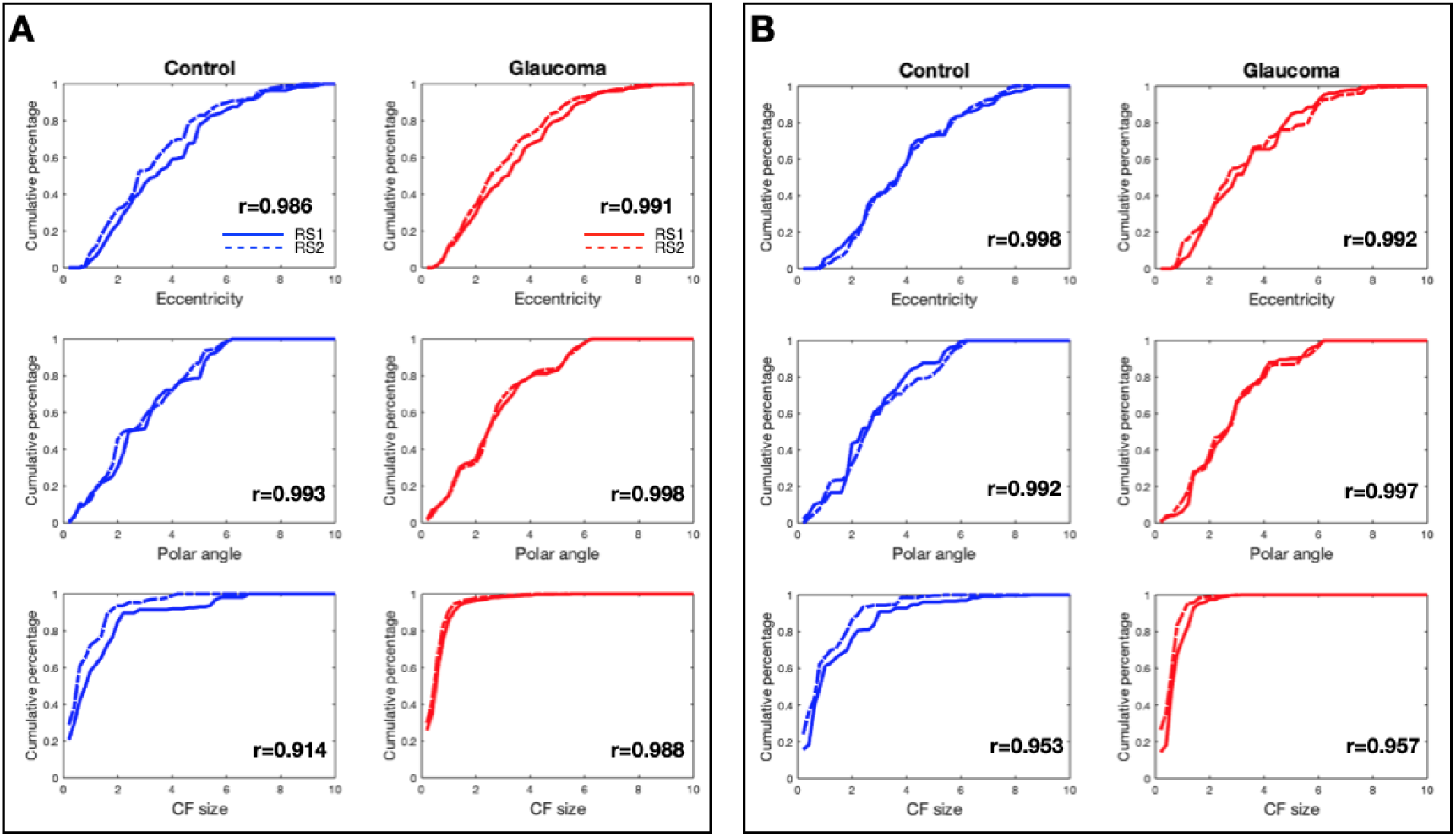
Test-retest evaluation of the RS scans. The median cumulative distributions of each CF parameter are plotted for RS1 (line) and RS2 (dashed line). Panel A shows results for V1>V2 while panel B shows those for V1>V3. Intraclass correlation coefficients (ICC) were calculated across all paired participants and for each ROI separately.

**Figure S6.**
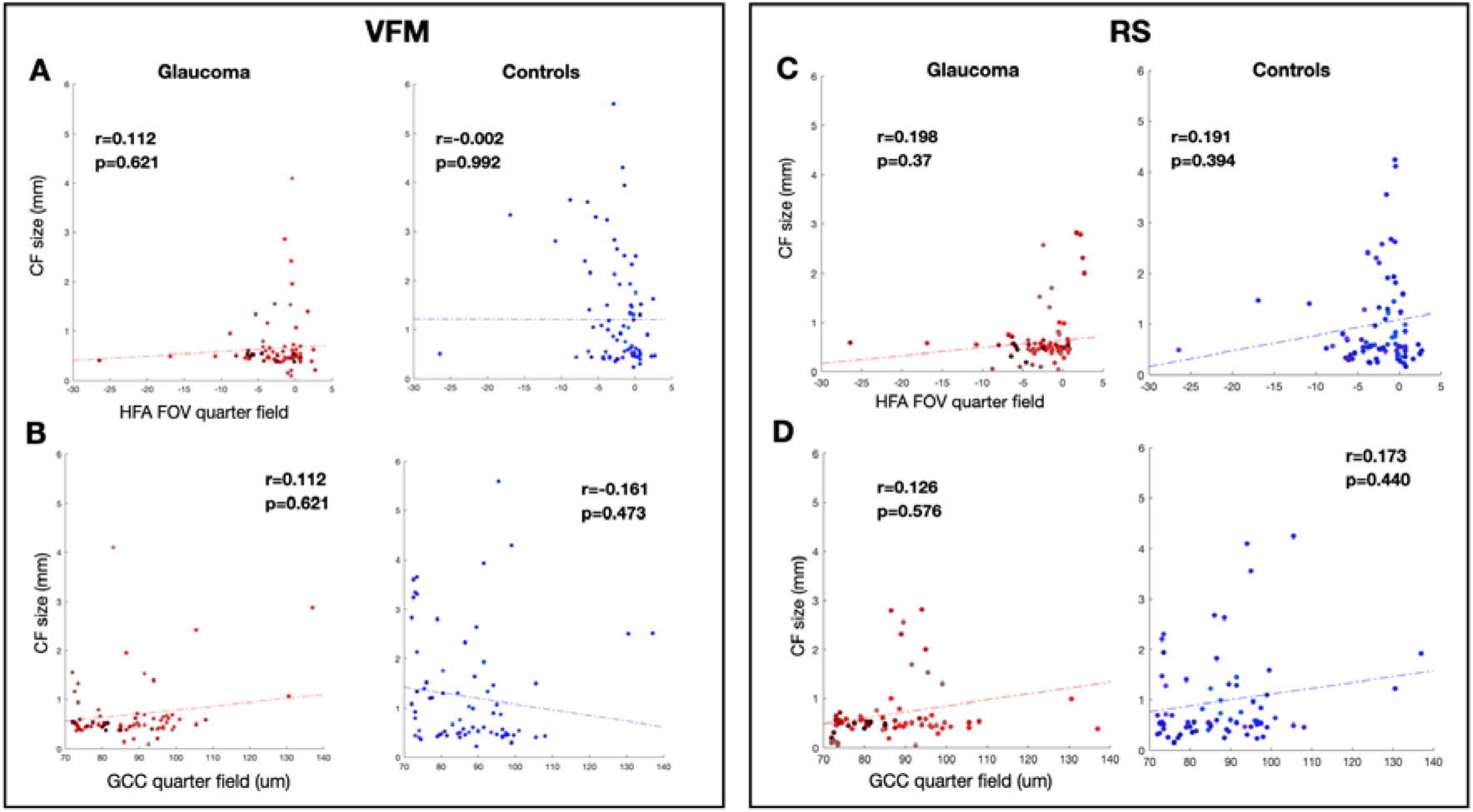
CF size as a function of glaucoma severity based on HFA and OCT data of V1>V3 area. Correlation of CF size obtained during VFM- and RS1 is correlated with with HFA and GCC from control and glaucoma participants. Each data point is from a separate quadrant of an individual participant. Panels A and C show the correlation of CF size with HFA, while Panels B and D show the correlation of CF size with GCC value, for VFM and RS respectively.

**Figure S7.**
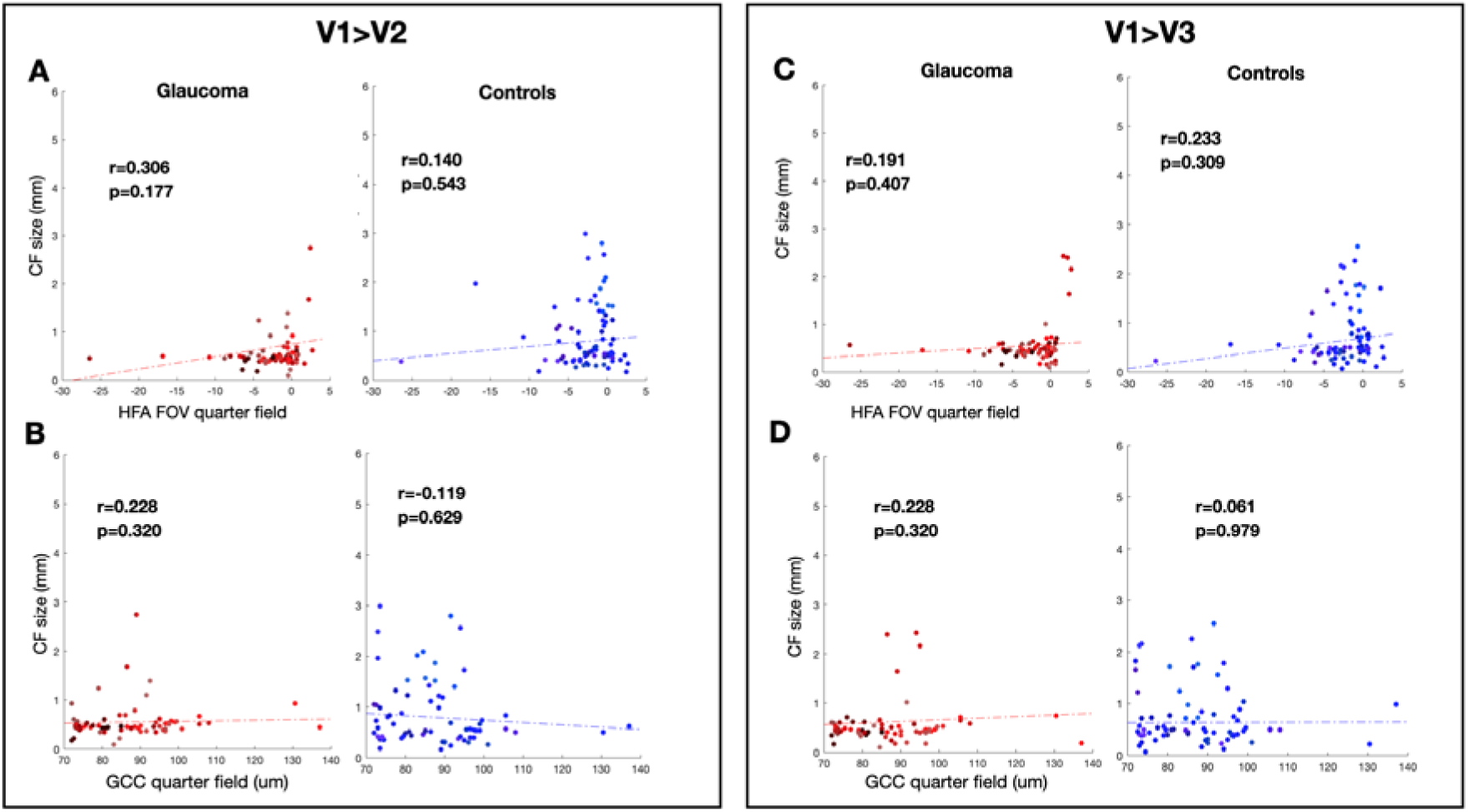
CF size for V1>V2 (left) and V1>V3 (right) obtained during RS (scan 2) as a function of glaucoma severity as based on either HFA or OCT. Each data point is from a separate quadrant of an individual participant. Panels A and C show the correlation of CF size with HFA, while Panels B and D show the correlation of CF size with GCC value, for VFM and RS respectively.

## Data Availability

All data produced in the present study are available upon reasonable request to the authors

